# Epidemiology and transmission dynamics of COVID-19 in two Indian states

**DOI:** 10.1101/2020.07.14.20153643

**Authors:** Ramanan Laxminarayan, Brian Wahl, Shankar Reddy Dudala, K. Gopal, Chandra Mohan, S. Neelima, K. S. Jawahar Reddy, J. Radhakrishnan, Joseph A. Lewnard

## Abstract

Although most COVID-19 cases have occurred in low-resource countries, there is scarce information on the epidemiology of the disease in such settings. Comprehensive SARS-CoV-2 testing and contact-tracing data from the Indian states of Tamil Nadu and Andhra Pradesh reveal stark contrasts from epidemics affecting high-income countries, with 92.1% of cases and 59.7% of deaths occurring among individuals <65 years old. The per-contact risk of infection is 9.0% (95% confidence interval: 7.5-10.5%) in the household and 2.6% (1.6-3.9%) in the community. Superspreading plays a prominent role in transmission, with 5.4% of cases accounting for 80% of infected contacts. The case-fatality ratio is 1.3% (1.0-1.6%), and median time-to-death is 5 days from testing. Primary data are urgently needed from low- and middle-income countries to guide locally-appropriate control measures.

---

Severe acute respiratory syndrome coronavirus 2 (SARS-CoV-2), the virus that causes coronavirus disease 2019 (COVID-19), has spread rapidly around the world since emerging in Wuhan, China in late 2019 (*1*). Current understanding of COVID-19 comes largely from disease surveillance and epidemiologic studies undertaken in early phases of the pandemic in China (*1*–*3*) and high-income countries of Europe (*4, 5*) and North America (*6*–*8*). However, most confirmed cases of COVID-19 have now occurred in low- and middle-income countries (LMICs), where a substantial proportion of individuals are at increased risk of severe outcomes and face barriers to accessing quality health services (*9*–*11*). While multiple modeling studies have sought to assess how COVID-19 might affect individuals and communities in such settings (*12*–*14*), almost no primary data about the transmission dynamics and clinical outcomes of COVID-19 in LMICs are available to validate these models and inform locally-appropriate intervention strategies (*15*).

Several considerations highlight the potential for differences in SARS-CoV-2 epidemiology in LMICs as compared to high-income settings. Transmission of the virus occurs directly from person-to-person through physical contact and through respiratory droplets and aerosols (*16, 17*). Inadequate housing quality and prevalent overcrowding may augment transmission risk among close contacts (*18, 19*), potentially impacting the effectiveness of stay-at-home orders and other mitigation measures that have achieved success in high-income countries (*20*). Enteric shedding of SARS-CoV-2 has further suggested the possibility of transmission via the fecal-oral route (*21*), which may be of particular importance in LMIC settings where many individuals lack sufficient access to clean water, handwashing facilities, and improved sanitation (*22*). Differences in demographic structure of LMICs and high-income settings may further influence patterns of spread as well as the distribution of population susceptibility (*23*). Intergenerational interactions, which are more common in many LMICs, may accelerate the epidemic, as has been observed in Italy where such interactions are also common (*23, 24*). While older adults have experienced the greatest burden of severe COVID-19 cases and deaths in high-income settings (*25*), high prevalence of chronic and acute comorbidities, and exposure to indoor and ambient air pollution, may exacerbate susceptibility within younger LMIC populations (*26*–*28*).

Over 1.3 billion people are at risk of SARS-CoV-2 infection in India, where concerns over COVID-19 have prompted large-scale containment strategies at the national, state, and local levels (*29*). The country’s first known COVID-19 case, documented on 30 January 2020, was an Indian national returning from China (*30*). In the initial weeks of the outbreak in India, transmission was facilitated by importation and travel-associated cases; however, national guidance aiming to prioritize testing for high-risk individuals might have precluded the detection of community transmission. Religious congregations, including a large gathering in Delhi in late March, served as super-spreading events after many attendees returned to their hometowns. Many states began implementing non-pharmaceutical interventions shortly thereafter to slow the spread of the disease, and a country-wide lockdown was initiated on 25 March (*29*).

Andhra Pradesh and Tamil Nadu are two states in the south of India that together account for approximately 10% of the country’s total population, with 127.8 million residents (**Fig 1A**). The two states are known for their public health capacity and effective primary healthcare delivery models (*31, 32*). Both states initiated rigorous disease surveillance and contact tracing early in response to the pandemic. Procedures include syndromic surveillance and SARS-CoV-2 testing for all individuals seeking care for severe acute respiratory illness or influenza-like illness at healthcare facilities; delineation of 5km “containment zones” surrounding cases for daily house-to-house surveillance to identify individuals with symptoms; and daily follow-up of all contacts of laboratory-confirmed or suspect COVID-19 cases, with the aim of testing these individuals 5-14 days after their contact with a primary case, irrespective of symptoms, to identify onward transmission (*33, 34*). We analyzed comprehensive testing and contact tracing data from these programs aiming to understand transmission dynamics and clinical outcomes of COVID-19 in South India, and to provide insights into control of SARS-CoV-2 in similar LMIC settings.

**Fig 1:**
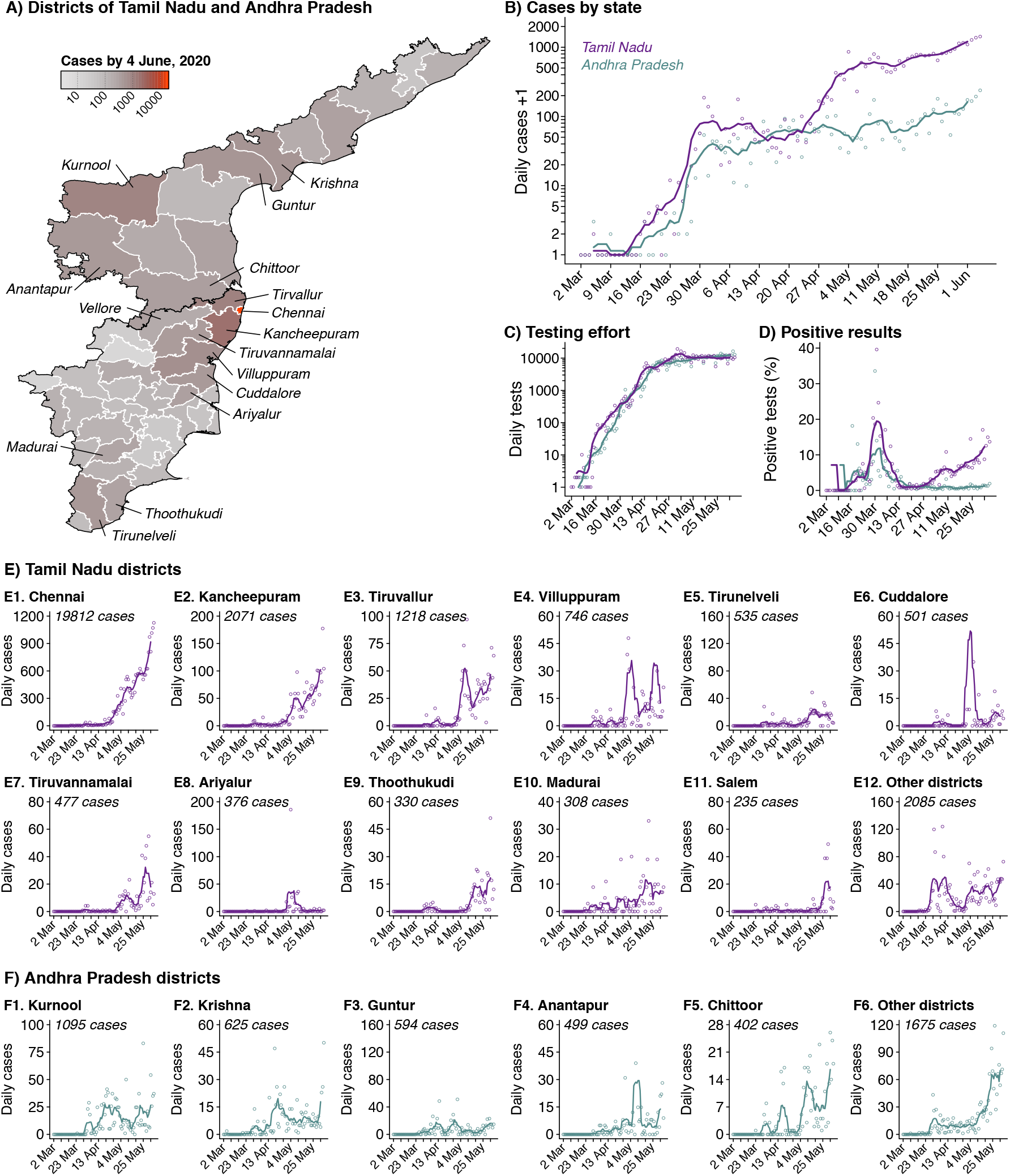
Tests performed and cases detected in Tamil Nadu and Andhra Pradesh, India. (**A**) We illustrate total cases by district within the two states; shading of regions on the choropleth map indicates higher case numbers. Districts are plotted according to 2019 administrative boundaries and do not reflect the recent bifurcation of Tirunelveli, Villuppuram, Vellore, and Chengalpattu districts. (**B**) We plot cases detected each day in each state (points) and 7-day moving averages (lines). Cases are aggregated by testing date. The same points and lines scheme applies to all panels of the figure; data are plotted in lavender and blue for Tamil Nadu and Andhra Pradesh, respectively. (**C**) We illustrate diagnostic tests conducted each day and (**D**) the proportion of tests yielding positive results. The high proportion of positive tests from late-March to mid-April, while case number remained relatively stable, may indicate a period during which cases were undercounted due to limited testing capacity. Last, we plot new incident cases by testing date for the districts of (**E**) Andhra Pradesh and (**F**) Tamil Nadu with the highest total case numbers.

## Disease surveillance

In India, surveillance of COVID-19 was initiated with airport screening for severe acute respiratory infection, especially for travelers from China. Tamil Nadu further instituted thermal and clinical screening at land borders with other states on 4 March. Nationwide, testing was initially prioritized for symptomatic individuals with history of travel or contact with a confirmed COVID-19 case within the previous 14 days, and was expanded to include all symptomatic individuals and asymptomatic contacts of confirmed cases in states between 20-28 March. We detail the timeline of changes in surveillance practices at federal and state levels in the **Materials and Methods**.

Tamil Nadu and Andhra Pradesh each recorded their first laboratory-confirmed COVID-19 cases on 5 March; our analyses include data collected through 4 June, at which time Tamil Nadu and Andhra Pradesh had conducted 518,935 and 476,988 diagnostic tests for SARS-CoV-2, respectively, identifying 28,694 positive cases in Tamil Nadu and 4,890 in Andhra Pradesh (**Fig. 1B**; **Table S1**). Numbers of new cases detected each day increased rapidly from 2 to 186 in Tamil Nadu and from 2 to 53 in Andhra Pradesh between 16 and 30 March (**Fig 1C; Table S2**). The proportion of tests yielding positive results peaked at 39.7% in Tamil Nadu and 33.5% in Andhra Pradesh on 30 and 31 March, respectively, when the daily number of tests performed was low in the two states (range: 379-469 tests). Throughout early April, increases in the number of tests performed daily coincided with a reduction in the proportion of tests yielding positive results, and with increases in the absolute number of cases identified each day (**Fig 1D**). This observation suggests cases might be under-counted in the early phase of the epidemic in each state, due to limitations in testing availability and testing algorithms. Test positivity increased in Tamil Nadu after 13 April in connection with exponential growth in new reported cases, but remained low in Andhra Pradesh. By 4 June, Tamil Nadu and Andhra Pradesh identified 1,438 and 240 new cases among 10,485 and 12,845 tests performed, respectively.

An uneven geographic distribution of cases within these states reflects differing local epidemic trajectories (**Fig. 1E-F**). The district of Chennai, home to 5.0 million of the 77.6 million individuals residing in Tamil Nadu, accounted for 69.0% (*n*=19,812) of all cases in the state; an additional 11.5% (*n*=3289) of all cases occurred in immediately-adjacent districts. Whereas incidence followed a similar pattern of sustained increases in these districts from mid-April through early June, the adjoining districts of Cuddalore, Ariyalur, and Villuppuram experienced a short-lived outbreak brought under control in mid-May. In contrast, the outbreak in Andhra Pradesh has not been centralized. The Kurnool district, where 4.1 million of the 50.3 million inhabitants of Andhra Pradesh live, experienced 22.4% of all reported cases by 4 June.

### Reproductive numbers

We used the method of Cori and colleagues (*35*) to estimate the time-varying reproductive number *R*_*t*_, describing the number of secondary infections each infected individual would be expected to generate as a moving average over 7-day time windows for each state and district. Over the period of 22-25 March, *R*_*t*_ was estimated to range between 1.86 (95% confidence interval: 1.40-2.38) and 3.80 (3.22-4.45) in Tamil Nadu. Estimates of *R*_*t*_ were sustained below 1 by the end of India’s country-wide lockdown, ranging from 0.82 (0.75-0.90) to 0.92 (0.83-1.02; **Fig. 2A**) over the period of 10-16 April. Increases occurring thereafter have held *R*_*t*_ point estimates between 1.0-1.2, with the exception of a transient peak around 28 April when *R*_*t*_ reached 1.98 (1.83-2.15). This peak corresponds to increases in transmission intensity in multiple districts, with Tiruvallur and Villuppuram experiencing *R*_*t*_ as high as 3.02 (2.37-3.76) and 4.51 (3.30-5.91), respectively. Following an initial estimate of *R*_*t*_ of 2.03 (1.61-2.49) as of 26 March, Andhra Pradesh saw a steady reduction in transmission intensity, with *R*_*t*_ reaching 0.93 (0.82-1.06) as of 5 April (**Fig. 2B**). Estimates thereafter have held steadily in the range of 1.0-1.2, with short periods around 28 April to 3 May and 11-15 May when *R*_*t*_ was sustained below 1. While *R*_*t*_ estimates are sensitive to changes in surveillance practices and superspreading events, particularly at early phases of the epidemic, these observations in Tamil Nadu and Andhra Pradesh broadly indicate a reduction in transmission following the implementation of multiple nonpharmaceutical interventions during late March and April (**Fig. 2C**), including the country-wide lockdown, closure of schools and nonessential businesses, and issuance of local stay-at-home and social distancing orders.

**Fig 2:**
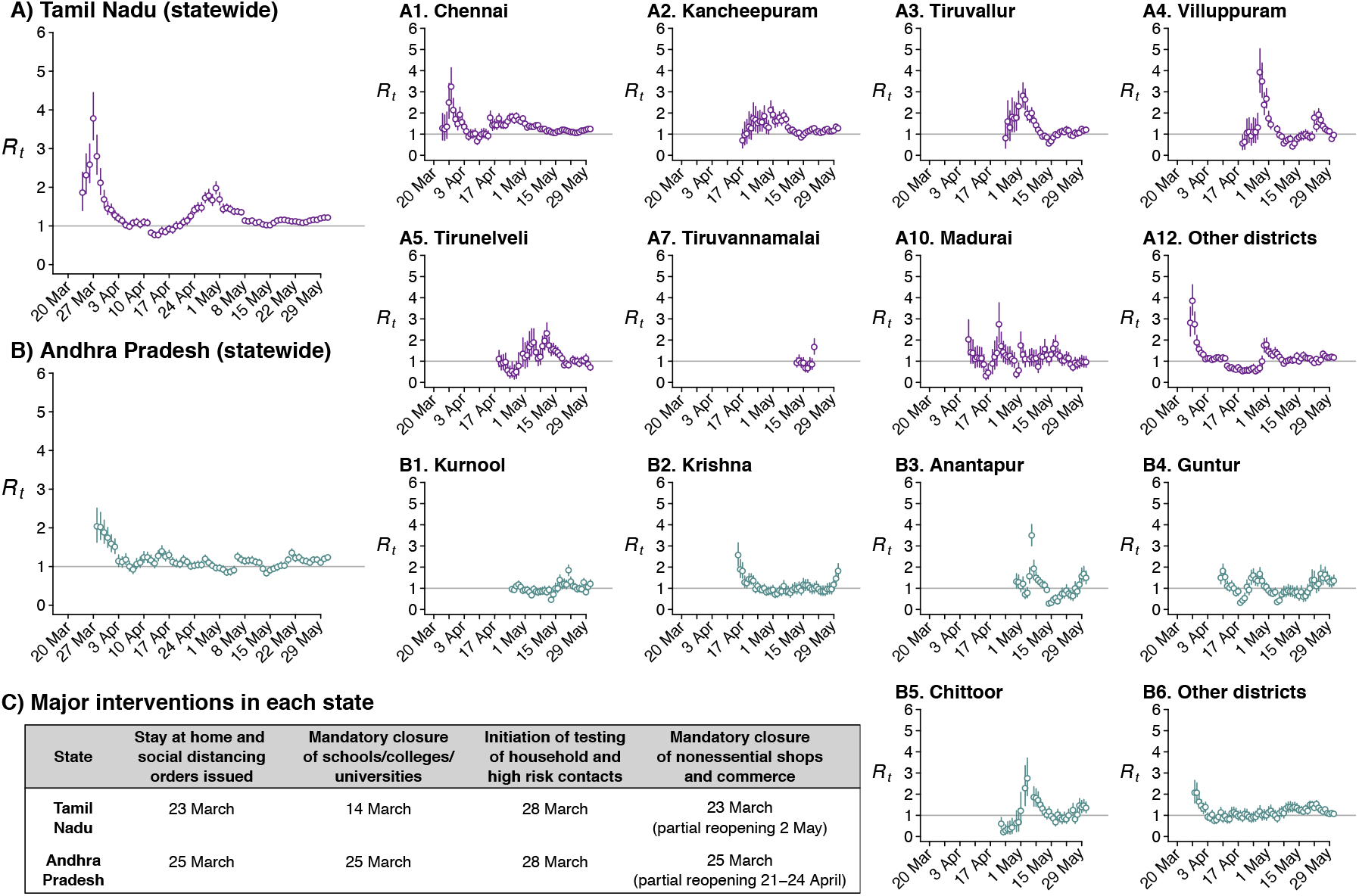
Instantaneous reproductive number plotted by state and district. We plot state-level and district-level estimates of the instantaneous reproductive number, *R*_*t*_, for cases tested on each day *t*, for (**A**) Tamil Nadu and (**B**) Andhra Pradesh. Lines around point estimates signify 95% credible intervals. (**C**) We indicate dates of major public interventions undertaken in each state.

### Contact tracing

Data were available from 64,031 contacts of 4,206 confirmed cases who were tested by 4 June in Tamil Nadu and by 29 May in Andhra Pradesh (**Table S3**). Up to 951 contacts were traced for each index case and tested within 5-14 days of exposure according to contact tracing protocols; 11 cases were linked to ≥200 contacts, whereas 50% of cases for whom contact tracing data were available had 7 or fewer contacts traced (**Fig. 3A**). No positive contacts were identified for 82.7% of index cases for whom contact-tracing data were available (**Fig. 3B**). In total, 5.4% of index cases accounted for 80% of positive contacts, and 1.5% of index cases accounted for 50% of positive contacts, suggesting a predominant role of super-spreading events in SARS-CoV-2 transmission in this setting (*36*). Fitting a negative binomial distribution to the number of infected contacts linked to each index case, we estimated a dispersion parameter of 0.072 (0.065-0.079). This estimate falls within the range estimated for SARS-CoV-2 outbreaks in other settings (*37*), and closely resembles estimates from prior outbreaks of severe acute respiratory syndrome (SARS), measles, and smallpox (*38*).

**Fig 3:**
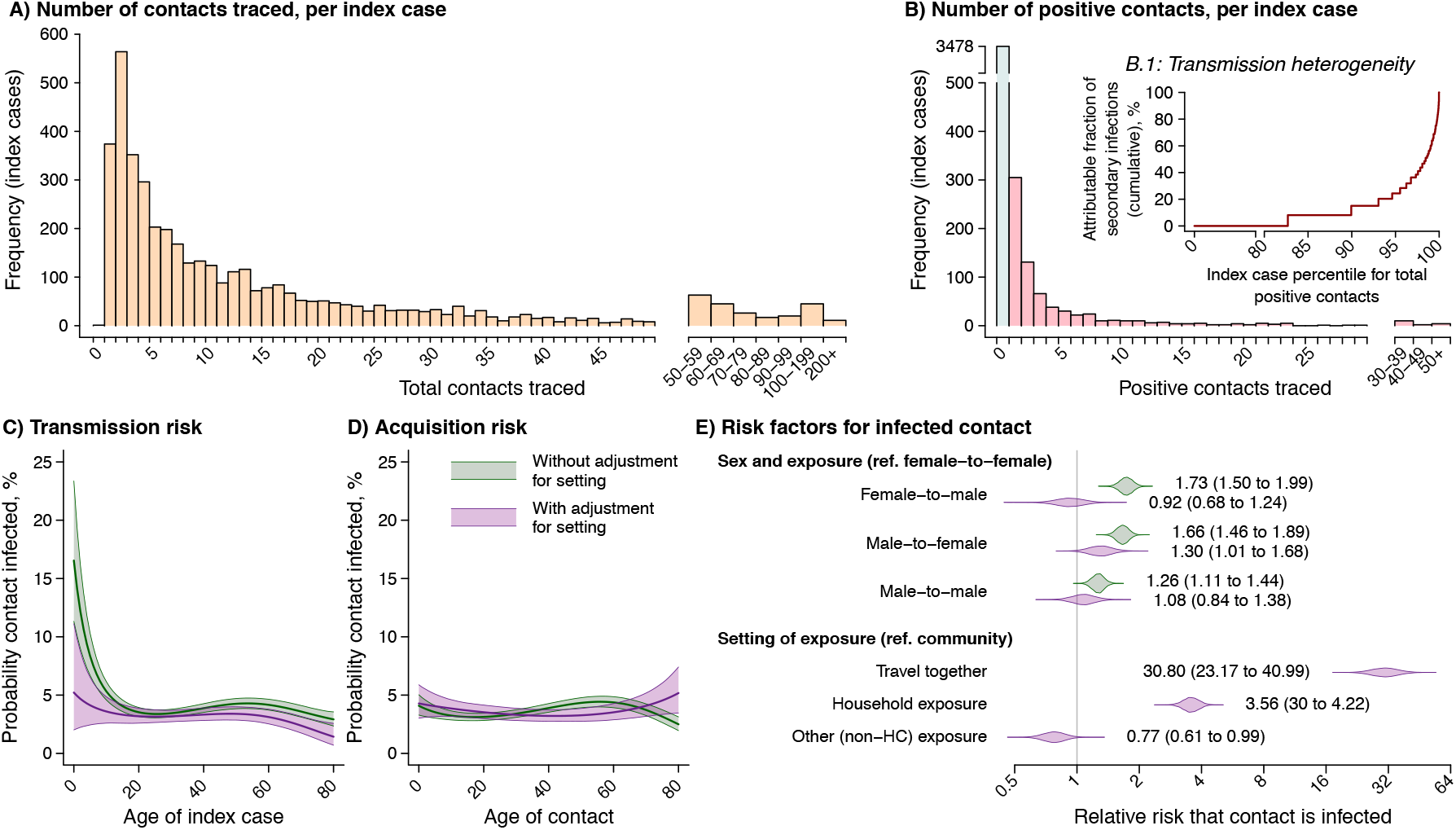
Contact tracing and infection outcomes. (**A**) We plot the distribution of the number of contacts traced for each index case in Tamil Nadu and Andhra Pradesh, binning values ≥50. (**B**) We illustrate the number of positive contacts traced from each index case, and (inset panel) the cumulative attributable proportion of secondary infections (y-axis) associated with each quantile value (x-axis) for the number of positive contacts traced per index case. (**C**) We illustrate risk of transmission as a function of index case age, for a male index case and a 35-year old male contact; shaded areas indicate 95% confidence bounds around point estimates (lines). We plot estimates based on models with (in violet) and without (in green) adjustment for transmission setting. (**D**) We illustrate infection risk for a secondary contact as a function of age, assuming a male contact and a 35-year old male index case. (**E**) We illustrate estimated effect sizes for predictors of a positive test result, addressing (top) sex of the index case and contact, and (bottom) exposure setting. Shaded areas indicate probability density for each estimate, with (violet) and without (green) adjustment for exposure setting, consistent with the left panel. Numbers in parentheses indicate 95% confidence intervals around point estimates of the adjusted relative risk.

Assuming test-positive contacts were infected by the index case to whom they were traced, we estimated that the overall secondary attack rate (or risk of transmission from an index case to a contact) was 6.0% (95% confidence interval: 5.0-7.3%). Data on contact settings, available for 18,485 contacts of 1,343 index cases, revealed considerable differences in transmission risk associated with differing types of interaction (**Fig. 3E**; **Table S4**-**S6**). Secondary attack rate estimates ranged from 1.0% (0.0-5.4%) in healthcare settings to 2.6% (1.6-3.9%) in the community and 9.0% (7.5-10.5%) in the household; in total, 48.3% of all positive contacts were traced to an index case in their household. In comparison to individuals who came into contact with an index case in the community, those who shared a household with the index case were 3.56 (2.99-4.22) times more likely to test positive, after adjusting for other risk factors (**Table S5**). Individuals who traveled in close (<1m) proximity to an index case in a shared conveyance were at the greatest risk of infection: this exposure was associated with 30.61 (23.03-40.75) fold higher risk than community exposure. These findings are consistent with studies from other countries that demonstrate households and transportation as high-risk exposure settings (*39*–*43*).

The probability for contacts to have a positive test result was not clearly associated with their age (**Fig. 3D**). While contacts of index cases who were children appeared more likely to be infected than contacts of adult index cases, this pattern did not persist after adjusting for the fact that contact with children more often occurred in household settings (**Fig. 3C**; **Tables S4**-**S5**). A similar finding emerged when assessing transmission risk as a function of the sex of index cases and their contacts (**Fig. 3E**; **Tables S4**-**S5**). Prior to adjustment for contact setting, mixed-sex interactions appeared to be associated with higher risk of transmission. Male contacts of female index cases were 36.9% (24.2-51.1%) more likely to test positive than male contacts of male index cases, while female contacts of male index cases were 66.1% (45.6-89.1%) more likely than female contacts of female index cases to test positive. Accounting for contact setting attenuated the strength of these associations. Following adjustment, we estimated that male index cases had 30.0% (0.8-66.5%) higher risk of transmitting to female contacts in comparison to female index cases, whereas differences in risk of transmission to male contacts from male and female index cases were not statistically meaningful.

### Mortality among COVID-19 cases

Analyses of a sub-cohort of 5,394 cases in Tamil Nadu and 1,939 cases in Andhra Pradesh who tested positive at least 30 days before the end of the study follow-up period provided an opportunity to estimate case-fatality ratios and time-to-death. All suspect and confirmed COVID-19 cases in India, including those comprising the surveillance data in Tamil Nadu and Andhra Pradesh, were admitted to dedicated COVID-19 facilities, until 5 May and 27 April, respectively, when asymptomatic and pre-symptomatic individuals were allowed to isolate at home; as such, testing dates in this sample are comparable to admission dates in other settings with hospital-based isolation of COVID-19 cases. Overall, 1.3% (1.0-1.6%) of cases died within 30 days of testing. Recovery was less frequent among older cases in comparison with younger cases; within the sub-cohort, no deaths occurred at ages 0-17 years, and case fatality ratios were 0.13% (0.04-0.28%) at ages 18-39 years, 2.3% (1.8-2.9%) at ages 40-64 years, and 6.5% (4.4-8.9%) at ages ≥65 years. Within each age group, men had greater risk of mortality than women (**Fig. 4F-H**). Following testing, the median time-to-death among fatal cases was 5 days, and 50% of fatal cases died within 1-12 days (interquartile range). These estimates of time-to-death in Tamil Nadu and Andhra Pradesh are well below what has been observed internationally: in the United States, median time-to-death from the date of hospital admission was 13 days (*8*), and the World Health Organization estimated time to death following onset of symptoms could range from two to eight weeks based on data from China (*44*). Between-setting differences in patients’ health status, healthcare systems capacity, and approaches to extending end-of-life care may contribute to observed variations in time-to-death.

**Fig 4:**
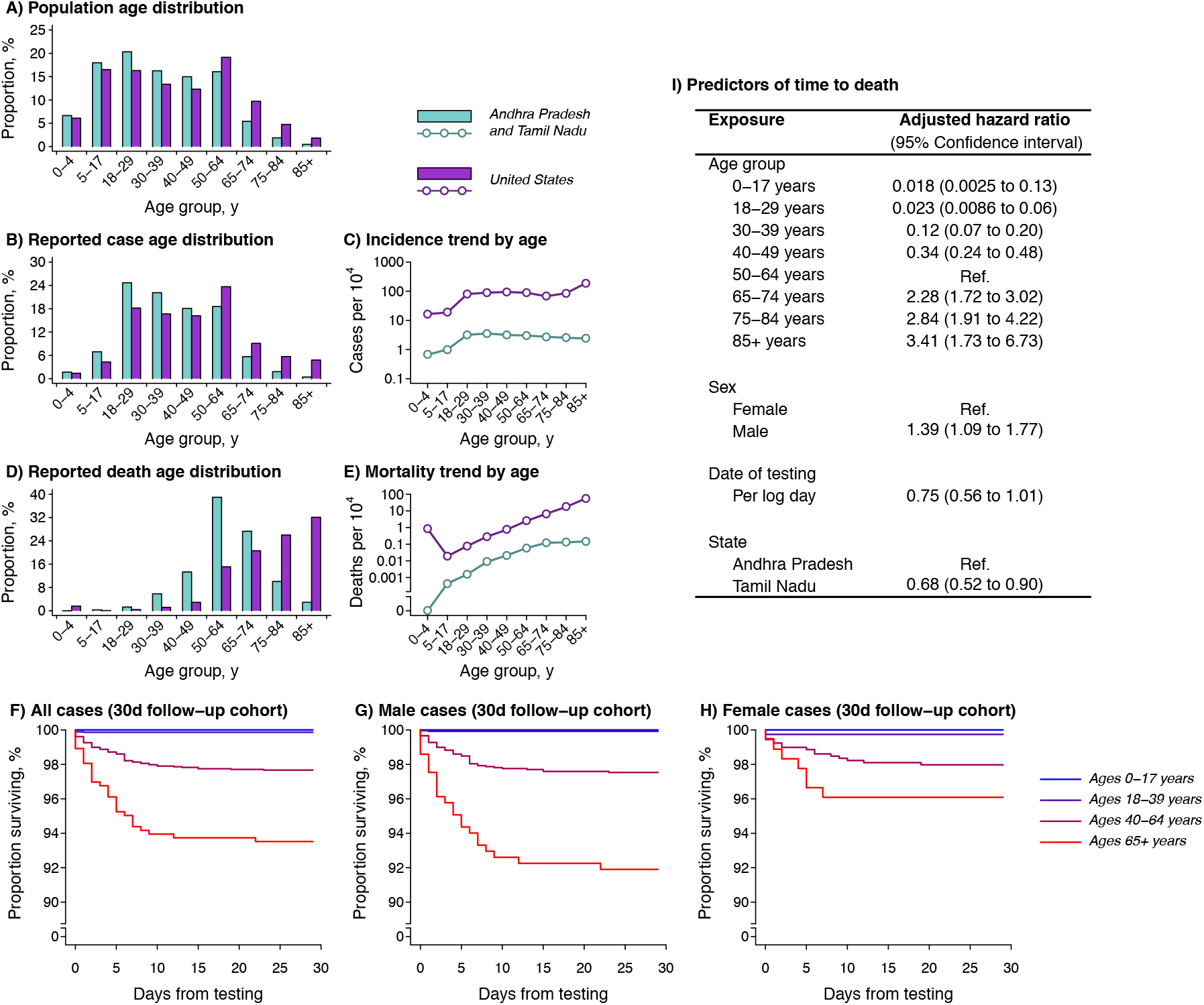
Age distribution and predictors of mortality. (**A**) We illustrate the age distribution of the population of Tamil Nadu and Andhra Pradesh (blue) against the age distribution of the US population (purple) for comparison; underlying data are presented in **Table S9**. Estimates are census extrapolations for the year 2020 in both settings; age bins were selected based on reporting of United States COVID-19 surveillance data. (**B**) We next illustrate the age distribution of cases and (**C**) cumulative incidence of COVID-19 by age in the two countries, and (**D**) the age distribution of deaths and (**E**) cumulative COVID-19 mortality by age. (**F**) Within a cohort of 7,322 cases tested ≥30 days before the last date of data capture, we plot survival by age (color ramp from blue, youngest, to red, oldest) over a 30-day period; we subdivide this cohort by sex with (**G**) males and (**H**) females. We aggregate age bins due to low counts within the 30 day follow-up cohort. (**I**) We indicate hazard ratios for the outcome of death by 4 June among all diagnosed COVID-19 cases in Tamil Nadu and Andhra Pradesh.

In a survival analysis of the full cohort of COVID-19 patients in Tamil Nadu and Andhra Pradesh, mortality by 4 June, 2020 was independently associated with older age, with stepwise increases in the adjusted hazard ratio of death for each successive age group (**Fig. 4I**; **Table S7**). Additional predictors of mortality included being male (adjusted hazard ratio: 1.39 [1.09-1.77] compared with being female), receipt of a test earlier in the epidemic (adjusted hazard ratio: 0.75 [0.56-1.01] for each log-day of testing from 1 March), and state of residence (adjusted hazard ratio: 0.68 [0.52-0.90] for residents of Tamil Nadu compared to Andhra Pradesh; **Fig. 4I**). Similar age and sex associations with mortality have been reported in many settings (*1*–*7, 45, 46*). The reduction in mortality over the course of the epidemic likely reflects strengthening of clinical care in both states, as well as increasing detection of less-severe cases as a result of expanded testing. Higher mortality among cases in Andhra Pradesh is noteworthy given the lower proportion of tests yielding positive results in this state. This finding may suggest healthcare or other factors in the relatively urbanized state of Tamil Nadu contribute to improved recovery among patients.

Among 280 decedents in the two Indian states for whom medical histories were available, the most prevalent comorbid conditions were diabetes (53.2%), sustained hypertension (51.8%), coronary artery disease (13.6%), and renal disease (10.4%; **Table S8**). Prevalence of comorbidities was higher among decedents at older ages (prevalence by age: 61.9%, 77.2%, and 83.3% at ages 0-39, 40-64, and ≥65 years). At least one comorbid condition was noted among 78.6% of fatalities, in comparison to 21.8% of fatalities in the United States as of 30 May, 2020 (*47*).

### Epidemiological comparison to high-income settings

The 33,584 confirmed cases in Tamil Nadu and Andhra Pradesh showed a younger age distribution than cases reported in the United States as of 27 June, 2020 (**Fig. 4A-C** (*48*)). Comparing cumulative COVID-19 incidence across ages revealed the observed differences surpassed expectations based on population age distributions alone (**Table S9**). Although lower across all age groups in Tamil Nadu and Andhra Pradesh in comparison to the United States, age-specific COVID-19 incidence increased sharply in both settings between the 5-17 year and 18-29 year age groups. Whereas incidence declined steadily at ages older than 30-39 years in the two Indian states, incidence increased at ages ≥65 years in the United States. We further address this observation in the **Supplementary text**.

In the two Indian states, only 13.0% of 308 COVID-19 deaths occurring on or before 4 June, 2020 were among individuals ages ≥75 years, compared with 58.1% of COVID-19 deaths in the United States (**Fig. 4D**; **Tables S7**-**S9**). Age-specific COVID-19 mortality was lower in Tamil Nadu and Andhra Pradesh compared with the United States, consistent with the lower reported incidence of disease. While COVID-19 mortality trended upward across ages in the two Indian states, mortality plateaued at ages ≥65 years, in contrast to observations in the United States where COVID-19 mortality reached 55.9 deaths per 10,000 individuals ages ≥85 years; this observation was consistent with the relatively lower incidence of disease at the oldest ages within the two Indian states. Additionally, no deaths were observed among 564 reported cases affecting children ages 0-4 years in Tamil Nadu and Andhra Pradesh, whereas the United States reported 0.8 COVID-19 deaths per 10,000 children in this age group (**Fig. 4E**).

## Discussion

This analysis provides insights into the epidemiology and transmission dynamics of COVID-19 in resource-limited populations based on comprehensive surveillance and contact tracing data from the Indian states of Tamil Nadu and Andhra Pradesh. We estimated reproduction numbers in the range of ∼2-3 in these settings as of late March, 2020, and identified reductions in *R*_*t*_ associated with the implementation of India’s country-wide lockdown and social distancing interventions at state and local levels. In the largest prospective study to date on the risk of SARS-CoV-2 infection among exposed contacts of cases, we identify substantial variation in individuals’ likelihood of transmitting: no secondary infections were linked to 83% of cases whose contacts were traced. Our analysis also highlights important differences related to sources of transmission in Tamil Nadu and Andhra Pradesh. After adjusting for contact setting, we identify no difference in infection risk among contacts of infected children and adults. While the role of children in transmission has been debated (*49, 50*), our observation of equivalent transmission risk across ages suggests a prominent role of children and young adults in sustaining epidemics in this setting, where 33% of cases were <30 years old. Last, our analyses of mortality identify substantially shorter time to death than what has been reported in other settings (*51*), with 50% of fatalities occurring within the first 5 days of testing. This observation should inform efforts to address clinical demand as the pandemic continues to strain India’s healthcare system.

Our study has several strengths. Prospective testing of a large sample of exposed individuals through integrated active surveillance and public health interventions in Tamil Nadu and Andhra Pradesh provided an opportunity to characterize secondary attack rates, identify risk factors for transmission, and account for deaths outside of healthcare settings—a limitation of mortality surveillance in other settings (*45, 52, 53*). Comprehensive testing data further provided insight into how changes in case ascertainment may have impacted epidemiologic surveillance. However, several limitations should be considered. Contact tracing data were only available for 12.5% of all cases identified in the two states through 4 June. It is not feasible to identify every contact of a known case and efforts are likely biased toward close contacts. This limitation likely contributes to an underestimate of the true community secondary attack rate, and an overestimate of the proportion of exposures occurring in household settings. Another limitation was the lack of data on timing of exposure and symptoms onset in relation to testing dates; this necessitated assumptions about identification of true index cases. More robust temporal data would reduce the dependence on such assumptions, provide greater insight into the directionality of transmission, and reduce risk for misclassification of infection status among contacts with positive or negative RT-PCR results at the time of testing (*40, 54*). The lack of temporal data also prevented us from estimating several epidemiologic parameters of interest. Current estimates of both the incubation period (c. 4-6 days) period and the serial interval (c. 3-5 days) come from China (*1, 39, 55*–*58*). Several factors can modify the incubation period of respiratory viral infections, including the route of acquisition, the infectious dose, and the period of exposure to infected cases (*59*). The serial interval between infection or case onset times is expected to be lower in high-transmission settings. Data allowing estimation of these parameters for SARS-CoV-2 in LMICs are needed to inform quarantine policies and other epidemic response efforts. Last, some true positive cases might have been misclassified due to the imperfect sensitivity of RT-PCR for detecting SARS-CoV-2 infection, including among contacts tested as few as 5 days after exposure to a known case. Imperfect sensitivity has been attributed to inadequate sample collection procedures and low viral load in the nasopharynx, particularly for pre-symptomatic or asymptomatic cases (*60*). This limitation could lead to an overall underestimate of transmission.

Testing and contact tracing are critical components of an effective public health response to COVID-19 (*61, 62*). In our study, data generated by these activities within two states of South India provided key insights into the local epidemiology and transmission dynamics SARS-CoV-2, without competing with emergency response activities for limited resources: a high priority in many LMICs where health workers and diagnostic equipment are already in short supply (*15*). Similar studies are necessary to inform the successful adaption of epidemic control measures in low-resource settings globally.

## Data Availability

Aggregate data underlying the analyses are presented in the supplemental tables.

## Acknowledgments

We are indebted to the work of the Governments of Tamil Nadu and Andhra Pradesh as well as healthcare workers and field workers engaged in outbreak response in these settings.

## Author contributions

Conceptualization: RL, BW, JAL; Methodology: BW, JAL; Software: JAL; Formal analysis: JAL; Investigation: SRD, KG, CM, SN, KSJR, JR; Resources: SRD, KG, CM, SN, KSJR, JR; Data curation: RL, JAL; Writing – Original Draft: BW, JAL; Writing – Review & Editing: RL, BW, SRD, KG, CM, SN, KSJR, JR, JAL; Visualization: JAL.

## Supplementary Materials

### Materials and Methods

#### Epidemiological surveillance

Data for the analyses were generated through public health surveillance activities undertaken by the Health and Family Welfare Department of the Government of Tamil Nadu and the Department of Health, Medical and Family Welfare Department of the Government of Andhra Pradesh, in accordance with national and state policies. In India, COVID-19 surveillance was initiated on 7 February, 2020 with airport-based thermal screening for international travelers from affected countries. Traveler screening was expanded to include thermal screening of maritime passengers and crew members on 10 March, 2020. Intensified health screening at land borders was commenced on 15 March, 2020.

Testing guidelines changed several times throughout the epidemic in India. Beginning on 9 March, 2020, symptomatic individuals developing symptoms 14 days after close contact with a laboratory-confirmed case or travel history to COVID-19 affected countries within 14 days of the onset of symptoms were eligible for testing. Testing was expanded to all symptomatic individuals with international travel history, and to symptomatic healthcare workers managing patients with respiratory distress or severe acute respiratory infection, on 17 March, 2020. As of 20 March, 2020, testing indications were expanded to all symptomatic healthcare workers, all hospitalized patients with severe acute respiratory infection, and all asymptomatic direct and high-risk contacts of confirmed cases, who were recommended to receive one test within 5-14 days of contact with a confirmed case.

Health is decentralized in India and states are empowered to implement national programs and guidelines with consideration for local contexts. During the epidemic, states could intensify response measures but were not sanctioned to dilute or weaken directives from the central government. Tamil Nadu commenced airport-based screening for severe acute respiratory infection among incoming international travelers on 20 January, 2020 and initiated thermal and clinical screening at borders with the states of Kerala, Karnataka, and Andhra Pradesh as of 4 March, 2020. Beginning on 28 March, 2020 in Tamil Nadu, all individuals with travel history were screened for symptoms and kept under home quarantine for 21 days with health monitoring to determine disease onset.

Active house-to-house surveillance for influenza-like-illness was carried out by community health workers in “containment zones” (i.e., areas within 5km radius around case residences) in both Tamil Nadu and Andhra Pradesh beginning on 28 March, 2020. Community health workers visited households to identify suspected cases and identify the contacts of all suspected or confirmed cases. Information on all cases and their contacts was communicated daily to supervisory medical officers. The contacts of all cases were monitored daily for development of symptoms over 28 days in Tamil Nadu and for the longer of either 28 days, or 21 days from the last occurrence of a positive case, in Andhra Pradesh.

Testing was conducted via real-time polymerase chain reaction (RT-PCR) by both government and private labs, with positive, negative, and indeterminate results notified directly to the Health and Family Welfare Department of the Government of Tamil Nadu and the Department of Health, Medical and Family Welfare of the Government of Andhra Pradesh. Information reported for each case included the laboratory where testing was conducted, the date the sample was obtained, and the age and sex of the patient. Positive cases were issued unique state-assigned case identification numbers, including in the scenario of posthumous SARS-CoV-2 detection for individuals who died shortly after admission to hospitals, or who died in the community and had specimens collected for SARS-CoV-2 testing. Retrospective medical record reviews were undertaken for fatal cases; consistent with US studies (*47*), comorbidities were considered to include any history of diabetes mellitus, sustained hypertension, coronary artery disease, renal disease, chronic obstructive pulmonary disease, asthma, cancer, pulmonary or extrapulmonary tuberculosis, stroke, neurological or neurodevelopmental disorders, or liver disease.

Our analyses are limited to tests performed for the purpose of diagnosis. Tests undertaken to determine recovery after infection (for the purposes of hospital discharge) were not included. For individuals who received multiple tests during the course of the epidemic, our analysis includes all tests conducted up to and including the test yielding a positive result, resulting in the individual being reported as a confirmed COVID-19 case and assigned a unique case identification number.

#### Reproductive number estimation

We used the EpiEstim package in R (*35*) to estimate the instantaneous reproduction number *R*_*t*_. As neither the delay distribution nor its mean were known for either the time from infection to testing or from symptoms onset to testing, we defined *R*_*t*_ for the testing day *t*, recognizing that this value may thus provide a retrospective view of transmission conditions. Based on maximum-likelihood fits to data from a previous review (*57*), we specified hyperparameters for the mean and standard deviation of the serial interval distribution as Norm(*μ*_Mean_ = 3.96, *σ*_Mean_ = 0.22) and Norm(*μ* _SD_ = 4.76, *σ* _SD_ = 0.16), respectively, and estimated *R*_*t*_ over sliding 7-day windows based on vectors of daily case counts. We limited analyses to 7-day periods with ≥14 cases to ensure stability of estimates (**Fig. 2**).

#### Inferring risk factors for infection among contacts

We fit Poisson regression models to data on contacts’ recorded infection status and attributes of each contact and their respective index cases to estimate adjusted relative risks for infection of contacts as a function of these exposures (*63*). We conducted forward-and-backward variable selection to determine whether addition of parameters to the regression model (and subsequent deletion of included parameters, with each addition) resulted in improvements in fit. We penalized for model degrees of freedom via the Bayesian Information Criterion. We assessed models that included up to fourth-degree polynomial terms defined from the ages of index cases and their contacts, the sex of index cases and their contacts (and interactions between these), state (Tamil Nadu or Andhra Pradesh), and the date of testing for the index case. Because contact type data were available only from Tamil Nadu (resulting in a change in sample size when included or excluded from the model), this variable was left out of the selection procedure. The best-fit model identified by the variable selection procedure discarded the fourth-degree polynomial term for contact age and the testing date of the index contact. We compared adjusted relative risk estimates using this model against those obtained when accounting further for contact type in the Tamil Nadu data subset to assess residual confounding (**Fig. 3**; **Table S5**).

#### Secondary attack rate estimation

We computed secondary attack rates by dividing the number of positive contacts by the number of contacts traced, overall and for each reported exposure setting. We conducted statistical inference using the cluster bootstrap to resample index cases, summing positive contacts and total contacts among resampled index cases (**Table S6**).

#### Survival analysis

We estimated adjusted hazard ratios for the outcome of death on or before 4 June, 2020 using Cox proportional hazards models, accounting for age group, sex, state (Tamil Nadu or Andhra Pradesh), and testing date (log-transformed). We defined the 40-49-year-old age group as a reference when estimating hazard ratios for age-specific mortality, as it included the greatest number of patients. The day of testing provided the earliest anchoring point in our study due to a lack of information on symptoms onset or exposure dates, and is a close proxy for admission date as all COVID-19 cases were admitted to dedicated COVID-19 facilities, until 5 May (in Tamil Nadu) and 27 April (in Andhra Pradesh) when individuals without symptoms were permitted to isolate at home.

#### Supplementary text

##### Risk of infection and transmission among children

Our analyses do not identify strong evidence of differential risk of acquiring or transmitting infection across ages in Tamil Nadu and Andhra Pradesh, after accounting for the fact that many of the observed infectious contacts involving children occurred in household settings. Evidence of the role of children in transmission is inconsistent, and risk likely varies across locations in accordance with public health interventions including closure of school or daycare centers, stringency of stay-at-home orders, and aspects of the built environment of settings where contact with children occurs. Limited data on infection risk are available from prospective testing of exposed children, similar to our study. Two contact tracing studies in China yielded conflicting conclusions, with one reporting no difference in risk of acquiring infection across ages (although children were less likely to experience symptoms; (*39*)) while the other reported higher risk of infection at older ages (*40*). One study in Switzerland revealed adults served as index cases for a majority of familial transmission clusters (*64*), however it is difficult to determine the role of differential contact patterns among children and adults outside of the household in this context, particularly in light of the closure of schools and daycare facilities. While one modeling study estimated lower susceptibility among children than adults (*14*), this finding should be interpreted carefully: model-calibrated estimates of age-specific transmission rates may be confounded by differences in risk across the household, community, or other settings where age groups come into contact, and fitting to case-based surveillance data may render bias from under-ascertainment of pediatric infections that are less likely to present with severe symptoms (*65*).

Outside our study, we are not aware of estimates of the relative risk of transmission as a function of index case age from prospectively-followed contact pairs. The fact that children and adults shed similar viral loads when infected lends biological support to the plausibility of our findings of equal transmission risk after controlling for setting of exposure (*66*). Analyses of contact tracing datasets should be pursued with urgency to understand this aspect of transmission.

##### Differential incidence pattern by age in India and the United States

Several factors are likely to contribute to our observation of differential trends in COVID-19 incidence across older ages in the Indian states of Tamil Nadu and Andhra Pradesh compared to the United States and other high-income settings. While differences in surveillance cannot be excluded, the fact that older adults are more susceptible to severe disease, when infected, should lead to a consistent pattern of underestimating the prevalence of infection among younger age groups across settings. It is plausible that stringent stay-at-home orders for older Indian adults, coupled with delivery of essentials through social welfare programs and regular community health worker interactions, contributed to lower exposure to infection within this age group. An alternative explanation may relate to demographic and socioeconomic factors in the two settings. In comparison to 77 years in China, 79 years in the United States, and 83 years in Italy and South Korea, life expectancy at birth is 69 years in India (*67*). As such, socioeconomic factors distinguishing individuals who survive to ages ≥75 years or ≥85 years in India from the general population are likely more pronounced in India (*68*) than in higher-income settings with longer average life expectancies (*69*). This may exacerbate survival bias when comparing COVID-19 outcomes across ages in India, as higher socioeconomic status may predispose individuals to surviving to older ages while also contributing to lower risk of experiencing COVID-19 (*70*).

**Table S1:** Attributes of COVID-19 cases in Tamil Nadu and Andhra Pradesh.

| Attribute |  | Tamil Nadu, *n* (%) | Andhra Pradesh, *n* (%) |
| --- | --- | --- | --- |
| Total cases |  | 28,694 | 4,890 |
| Age | 0-4 years | 514 (2) | 50 (1) |
|  | 5-17 years | 2,072 (7) | 251 (5) |
|  | 18-29 years | 6,739 (23) | 1,548 (32) |
|  | 30-39 years | 6,324 (22) | 1,104 (23) |
|  | 40-49 years | 5,417 (19) | 655 (13) |
|  | 50-64 years | 5,466 (19) | 762 (16) |
|  | 65-74 years | 1,517 (5) | 384 (8) |
|  | 75-84 years | 500 (2) | 104 (2) |
|  | ≥85 years | 116 (<1) | 28 (1) |
|  | Unknown | 29 (<1) | 4 (<1) |
| Sex | Male | 17,796 (62) | 3,068 (63) |
|  | Female | 10,882 (38) | 1,797 (37) |
|  | Transgender | 16 (<1) | 1 (<1) |
|  | Unknown | 0 (0) | 24 (<1) |
| Testing date | Before 1 April, 2020 | 345 (1) | 144 (3) |
|  | 1-15 April, 2020 | 1,040 (4) | 541 (11) |
|  | 16-30 April, 2020 | 1,272 (4) | 936 (19) |
|  | 1-15 May, 2020 | 7,928 (28) | 1,008 (21) |
|  | 16-31 May, 2020 | 12,910 (45) | 1,486 (30) |
|  | 1-4 June, 2020 | 5,199 (18) | 775 (16) |

Values in the table present descriptive attributes of laboratory-confirmed COVID-19 cases in the two states.

**Table S2:** Population tested and test positivity in Tamil Nadu and Andhra Pradesh.

| Attribute | Tamil Nadu |  | Andhra Pradesh |  |
| --- | --- | --- | --- | --- |
|  | All tested, *n*<br>(%) | Positive, *n* (% of<br>those tested) | All tested, *n* (%) | Positive (% of those<br>tested) |
| Total tested | 518,935 | 28,694 (6) | 476,988 | 4,890 (1) |
| Age |  |  |  |  |
| 0-4 years | 11,255 (2) | 514 (5) | 4,809 (1) | 50 (1) |
| 5-17 years | 31,251 (6) | 2,072 (7) | 25,267 (5) | 251 (1) |
| 18-29 years | 177,395 (34) | 6,739 (4) | 144,952 (30) | 1,548 (1) |
| 30-39 years | 118,204 (23) | 6,324 (5) | 111,031 (23) | 1,104 (1) |
| 40-49 years | 77,830 (15) | 5,417 (7) | 76,475 (23) | 655 (1) |
| 50-64 years | 73,926 (14) | 5,466 (7) | 76,473 (16) | 762 (1) |
| 65-74 years | 18,524 (4) | 1,517 (8) | 28,544 (6) | 384 (1) |
| 75-84 years | 5,951 (1) | 500 (8) | 7,400 (2) | 104 (1) |
| ≥85 years | 1,355 (<1) | 166 (12) | 959 (<1) | 28 (3) |
| Unknown | 3,247 (1) | 29 (1) | 1083 (<1) | 4 (<1) |
| Sex |  |  |  |  |
| Male | 287,337 (55) | 17,796 (6) | 302,962 (64) | 3,068 (1) |
| Female | 231,333 (45) | 10,882 (5) | 173,832 (36) | 1,797 (1) |
| Transgender | 101 (<1) | 16 (16) | 112 (<1) | 1 (1) |
| Unknown | 164 (<1) | 0 (0) | 82 (<1) | 24 (29) |
| Testing date |  |  |  |  |
| Before 1 April, 2020 | 2,777 (1) | 345 (12) | 1,287 (<1) | 144 (11) |
| 1-15 April, 2020 | 31,701 (6) | 1,040 (3) | 17,276 (4) | 541 (3) |
| 16-30 April, 2020 | 103,669 (20) | 1,272 (1) | 98,856 (21) | 936 (1) |
| 1-15 May, 2020 | 176,352 (34) | 7,928 (4) | 132,378 (28) | 1,008 (1) |
| 16-31 May, 2020 | 162,052 (31) | 12,910 (8) | 174,037 (36) | 1,486 (1) |
| 1-4 June, 2020 | 42,377 (8) | 5,199 (12) | 53,154 (11) | 775 (1) |

Values in the table present descriptive attributes of all individuals who received real-time polymerase chain reaction (RT-PCR) tests in the two states.

**Table S3:** Attributes of cases and contacts in the contact-tracing dataset.

| Attribute | Tamil Nadu |  |  | Andhra Pradesh |  |  |
| --- | --- | --- | --- | --- | --- | --- |
|  | Index cases, *n*<br>(%) | All contacts, *n*<br>(%) | Positive contacts, *n*<br>(% of contacts tested) | Index cases, *n*<br>(%) | All contacts, *n*<br>(%) | Positive contacts, *n*<br>(% of contacts tested) |
| Total included | 1,351 | 18,813 | 789 | 2,855 | 45,218 | 3,004 |
| Age |  |  |  |  |  |  |
| 0-4 years | 14 (1) | 472 (3) | 30 (6) | 22 (1) | 600 (1) | 10 (2) |
| 5-17 years | 52 (4) | 1,951 (10) | 133 (7) | 200 (7) | 3,113 (7) | 260 (8) |
| 18-29 years | 330 (24) | 4,802 (26) | 184 (4) | 700 (25) | 8,005 (18) | 459 (6) |
| 30-39 years | 365 (27) | 3,772 (20) | 125 (3) | 658 (23) | 7,334 (16) | 600 (8) |
| 40-49 years | 300 (22) | 3,043 (16) | 131 (4) | 494 (17) | 5,303 (11) | 436 (8) |
| 50-64 years | 216 (16) | 2,821 (15) | 112 (4) | 545 (19) | 4,566 (10) | 567 (12) |
| 65-74 years | 50 (4) | 578 (3) | 34 (6) | 184 (6) | 1,638 (4) | 120 (7) |
| 75-84 years | 13 (1) | 173 (1) | 6 (3) | 43 (2) | 467 (1) | 51 (10) |
| ≥85 years | 4 (<1) | 41 (<1) | 4 (10) | 9 (<1) | 288 (1) | 0 (0) |
| Unknown | 7 (<1) | 1160 (6) | 30 (3) | 0 (0) | 13,904 (1) | 501 (4) |
| Sex |  |  |  |  |  |  |
| Male | 1,017 (75) | 10,151 (54) | 401 (4) | 1,816 (64) | 27,101 (60) | 1,786 (7) |
| Female | 332 (25) | 7,701 (41) | 382 (5) | 1,039 (36) | 18,117 (40) | 1,218 (7) |
| Transgender | 0 (0) | 0 (0) | 0 (Undef.) | 0 (0) | 0 (0) | 0 (Undef.) |
| Unknown | 2 (<1) | 961 (5) | 6 (1) | 0 (0) | 0 (0) | 0 (Undef.) |
| Contact setting |  |  |  |  |  |  |
| Community | 596 (34) | 9,628 (51) | 249 (3) | -- | -- | -- |
| Household | 997 (56) | 4,066 (22) | 380 (9) | -- | -- | -- |
| Travel together | 8 (<1) | 78 (<1) | 63 (81) | -- | -- | -- |
| Healthcare | 11 (1) | 210 (1) | 2 (1) | -- | -- | -- |
| Other | 151 (9) | 4,503 (24) | 92 (2) | -- | -- | -- |
| Not recorded | 12 (1) | 328 (2) | 3 (1) | 2,855 (100) | 45,218 (100) | 3,004 (7) |
| Testing date of index case |  |  |  |  |  |  |
| Before 1 April, 2020 | 116 (9) | 1,881 (10) | 56 (3) | 88 (3) | 3,290 (7) | 169 (5) |
| 1-15 April, 2020 | 226 (17) | 4,316 (23) | 90 (2) | 428 (15) | 12,245 (27) | 1,111 (9) |
| 16-30 April, 2020 | 270 (20) | 5,020 (27) | 132 (3) | 859 (30) | 12,142 (27) | 820 (7) |
| 1-15 May, 2020 | 599 (44) | 5,879 (31) | 243 (4) | 789 (28) | 11,402 (25) | 644 (6) |
| 16-31 May, 2020 | 138 (10) | 1,383 (7) | 237 (17) | 691 (24) | 6,139 (14) | 260 (4) |
| Not recorded | 2 (<1) | 334 (2) | 31 (9) | 0 (0) | 0 (0) | 0 (Undef.) |

Values in the table present descriptive attributes of laboratory-confirmed COVID-19 cases in the two states for whom contacts were traced, and descriptive attributes of contacts who were traced and who tested positive via real-time polymerase chain reaction (RT-PCR).

**Table S4:**
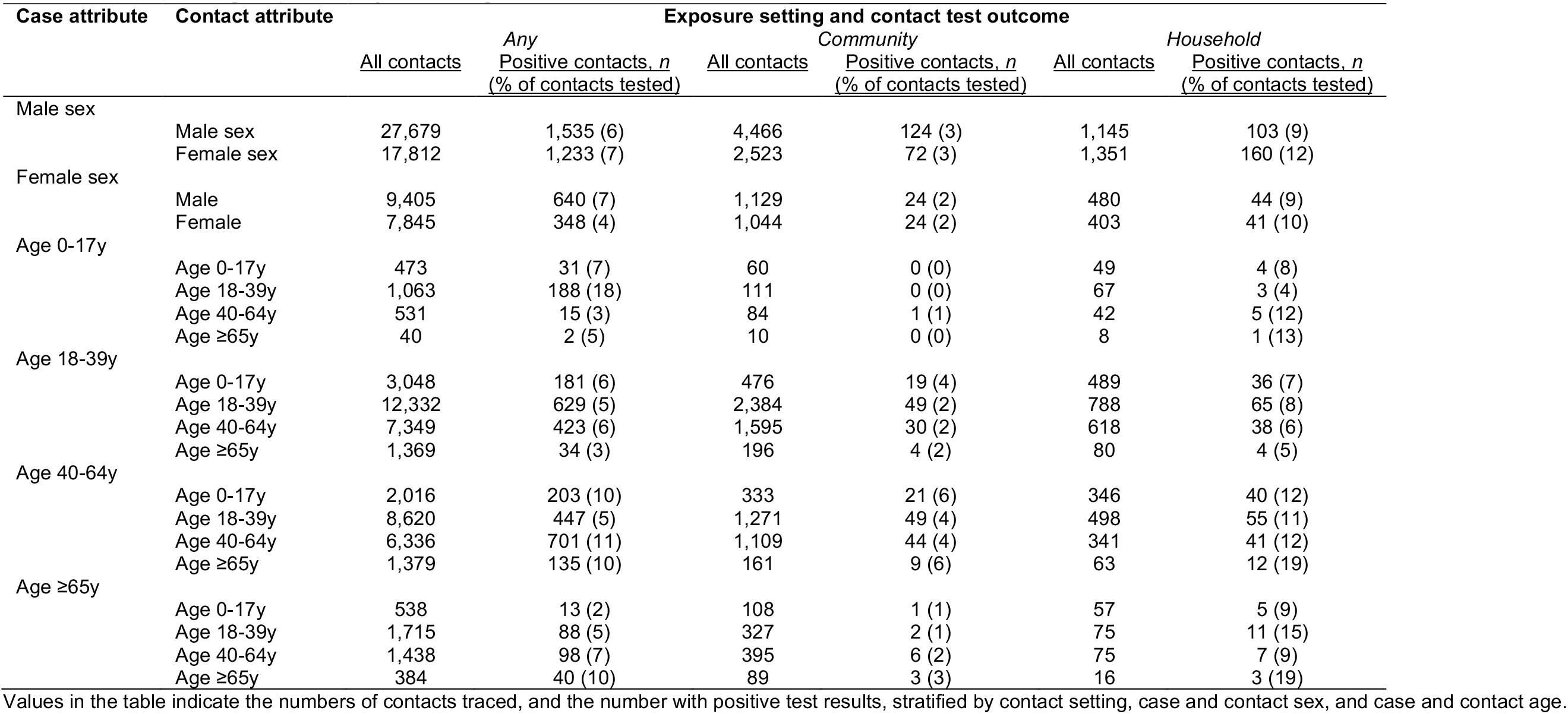
Contact setting frequencies by sex and age.

**Table S5:** Predictors of infection among contacts of known cases.

| Exposure |  | Adjusted relative risk (95% CI) | Adjusted relative risk, accounting for contact setting (95% CI) |
| --- | --- | --- | --- |
| Sexes | Female index, female contact | ref. | ref. |
|  | Female index, male contact | 1.73 (1.50, 1.99) | 0.92 (0.68, 1.24) |
|  | Male index, female contact | 1.66 (1.46, 1.89) | 1.30 (1.01, 1.68) |
|  | Male index, male contact | 1.26 (1.11, 1.44) | 1.08 (0.84, 1.38) |
| State | Andhra Pradesh | ref. | -- |
|  | Tamil Nadu | 0.53 (0.48, 0.57) | -- |
| Setting | Community | -- | ref. |
|  | Household | -- | 3.56 (3.00, 4.22) |
|  | Travel together | -- | 30.80 (23.17, 40.99) |
|  | Healthcare | -- | 0.36 (0.09, 1.44) |
|  | Other | -- | 0.77 (0.61, 0.99) |

**Table S6:** Secondary attack rates by setting.

| Setting | Estimate (95% CI) |
| --- | --- |
| Community | 2.6% (1.6-3.9%) |
| Household | 9.0% (7.5-10.5%) |
| Travel together | 80.8% (48.6-98.5%) |
| Healthcare | 1.0% (0.0-5.4%) |
| Other | 2.1% (0.4-4.4%) |
| Any setting | 6.0% (5.0-7.3%) |

Values in the table indicate the proportion of all traced contacts who test positive. We obtain confidence intervals via cluster-bootstrap resampling of index cases.

**Table S7:** Attributes of fatal COVID-19 cases in Tamil Nadu and Andhra Pradesh.

| Attribute |  | Tamil Nadu, *n* (%) | Andhra Pradesh, *n* (%) |
| --- | --- | --- | --- |
| Total deceased by 4 June, 2020 |  | 235 | 73 |
| Age | 0-4 years | 0 (0) | 0 (0) |
|  | 5-17 years | 1 (<1) | 0 (0) |
|  | 18-29 years | 3 (1) | 1 (1) |
|  | 30-39 years | 14 (6) | 4 (5) |
|  | 40-49 years | 30 (13) | 11 (15) |
|  | 50-64 years | 85 (36) | 35 (48) |
|  | 65-74 years | 69 (29) | 15 (21) |
|  | 75-84 years | 25 (11) | 6 (8) |
|  | ≥85 years | 8 (3) | 1 (1) |
|  | Unknown | 0 (0) | 0 (0) |
| Sex | Male | 162 (69) | 52 (71) |
|  | Female | 73 (31) | 21 (29) |
|  | Transgender | 0 (0) | 0 (0) |
|  | Unknown | 0 (0) | 0 (0) |

Values in the table present descriptive attributes of fatalities with SARS-CoV-2 detected via real-time polymerase chain reaction testing.

**Table S8:** Prevalence of comorbid conditions among fatal COVID-19 cases.

| Condition | Sex | Prevalence among decedents, % |  |  |  |
| --- | --- | --- | --- | --- | --- |
|  |  | 0-39y | 40-64y | ≥65y | All ages |
| All decedents with comorbidity data recorded | All | 21 | 145 | 114 | 280 |
|  | Males | 12 | 100 | 84 | 196 |
|  | Females | 9 | 45 | 30 | 84 |
| Diabetes mellitus | All | 4 (19) | 84 (58) | 61 (54) | 149 (53) |
|  | Males | 1 (8) | 60 (60) | 46 (55) | 107 (55) |
|  | Females | 3 (33) | 24 (53) | 15 (50) | 42 (50) |
| Sustained hypertension | All | 7 (33) | 72 (50) | 66 (58) | 145 (52) |
|  | Males | 5 (42) | 48 (48) | 45 (54) | 98 (50) |
|  | Females | 2 (22) | 24 (53) | 21 (70) | 47 (56) |
| Coronary artery disease | All | 0 (0) | 14 (10) | 24 (21) | 38 (14) |
|  | Males | 0 (0) | 9 (9) | 17 (20) | 26 (13) |
|  | Females | 0 (0) | 5 (11) | 7 (23) | 12 (14) |
| Renal disease | All | 2 (10) | 13 (9) | 14 (12) | 29 (10) |
|  | Males | 1 (8) | 9 (9) | 13 (15) | 23 (12) |
|  | Females | 1 (11) | 4 (9) | 1 (3) | 6 (7) |
| Chronic obstructive pulmonary disease | All | 0 (0) | 6 (4) | 12 (11) | 18 (6) |
|  | Males | 0 (0) | 4 (4) | 12 (14) | 16 (8) |
|  | Females | 0 (0) | 2 (4) | 0 (0) | 2 (2) |
| Asthma | All | 3 (14) | 6 (4) | 3 (3) | 12 (4) |
|  | Males | 0 (0) | 4 (4) | 1 (1) | 5 (3) |
|  | Females | 0 (0) | 2 (4) | 2 (7) | 7 (8) |
| Cancer | All | 1 (5) | 4 (3) | 7 (6) | 12 (4) |
|  | Males | 1 (8) | 2 (2) | 5 (6) | 8 (4) |
|  | Females | 0 (0) | 2 (4) | 2 (7) | 4 (5) |
| Pulmonary tuberculosis | All | 1 (5) | 3 (2) | 2 (2) | 6 (2) |
|  | Males | 1 (8) | 3 (3) | 2 (2) | 6 (3) |
|  | Females | 0 (0) | 0 (0) | 0 (0) | 0 (0) |
| Stroke | All | 0 (0) | 3 (2) | 4 (4) | 7 (2) |
|  | Males | 0 (0) | 2 (2) | 2 (2) | 4 (2) |
|  | Females | 0 (0) | 1 (2) | 2 (7) | 3 (3) |
| Neurological or neurodevelopmental disorder | All | 1 (5) | 1 (1) | 2 (2) | 4 (1) |
|  | Males | 0 (0) | 1 (1) | 0 (0) | 1 (1) |
|  | Females | 1 (11) | 0 (0) | 2 (7) | 3 (4) |
| Liver disease | All | 1 (5) | 0 (0) | 3 (3) | 4 (1) |
|  | Males | 1 (8) | 0 (0) | 2 (2) | 3 (2) |
|  | Females | 0 (0) | 0 (0) | 1 (3) | 1 (1) |
| Any comorbidity | All | 13 (62) | 112 (77) | 95 (83) | 220 (79) |
|  | Males | 7 (58) | 80 (80) | 68 (81) | 155 (79) |
|  | Females | 6 (67) | 32 (71) | 27 (90) | 65 (77) |
| Two or more comorbidities | All | 6 (29) | 71 (49) | 63 (55) | 140 (50) |
|  | Males | 3 (25) | 48 (48) | 45 (54) | 96 (49) |
|  | Females | 3 (33) | 23 (51) | 18 (60) | 44 (52) |
| Three or more comorbidities | All | 1 (5) | 17 (12) | 33 (29) | 51 (18) |
|  | Males | 0 (0) | 9 (9) | 26 (31) | 35 (18) |
|  | Females | 1 (11) | 8 (18) | 7 (23) | 16 (19) |

Values in the table indicate the number and proportion of decedents in each stratum with the listed comorbidities. Comorbidities data were unavailable for 28 decedents who are excluded from the table.

**Table S9:** Cumulative incidence of COVID-19 cases and mortality in Tamil Nadu, Andhra Pradesh, and the United States.

| Setting | Age group | Population at risk<br>(in 10,000s) | Cases<br>(in 10,000s) | Cumulative incidence<br>(per 10,000) | Incidence ratio<br>(ref. age 65-74y) | Deaths<br>(in 100s) | Cumulative mortality<br>(per 10,000) | Mortality ratio<br>(ref. age 65-74y) |
| --- | --- | --- | --- | --- | --- | --- | --- | --- |
| Tamil Nadu and Andhra Pradesh | 0-4 years | 849.7 | 0.1 | 0.7 | 0.24 | 0 | 0 | 0 |
|  | 5-17 years | 2298.0 | 0.2 | 1.0 | 0.37 | 0.01 | <0.01 | 0.0036 |
|  | 18-29 years | 2598.7 | 0.8 | 3.2 | 1.16 | 0.04 | 0.015 | 0.013 |
|  | 30-39 years | 2078.4 | 0.7 | 3.6 | 1.30 | 0.18 | 0.087 | 0.071 |
|  | 40-49 years | 1915.4 | 0.6 | 3.2 | 1.15 | 0.41 | 0.21 | 0.18 |
|  | 50-64 years | 2053.3 | 0.6 | 3.0 | 1.10 | 1.20 | 0.58 | 0.48 |
|  | 65-74 years | 692.6 | 0.2 | 2.7 | ref. | 0.84 | 1.21 | ref. |
|  | 75-84 years | 240.3 | 0.1 | 2.5 | 0.92 | 0.31 | 1.29 | 1.06 |
|  | ≥85 years | 60.9 | <0.1 | 2.4 | 0.87 | 0.09 | 1.48 | 1.22 |
| United States | 0-4 years | 1973.6 | 3.3 | 16.5 | 0.24 | 16.4 | 8.3 | 0.12 |
|  | 5-17 years | 5354.8 | 10.0 | 18.7 | 0.28 | 1.0 | 0.2 | 0.0029 |
|  | 18-29 years | 5287.1 | 42.3 | 80.0 | 1.19 | 4.1 | 0.8 | 0.012 |
|  | 30-39 years | 4337.5 | 38.8 | 89.5 | 1.33 | 12.3 | 2.8 | 0.042 |
|  | 40-49 years | 3992.9 | 37.7 | 94.3 | 1.40 | 29.8 | 7.5 | 0.11 |
|  | 50-64 years | 6211.0 | 55.1 | 88.7 | 1.32 | 154.9 | 24.9 | 0.37 |
|  | 65-74 years | 3148.7 | 21.2 | 67.2 | ref. | 211.4 | 67.1 | ref. |
|  | 75-84 years | 1540.7 | 13.3 | 86.0 | 1.28 | 266.8 | 173.1 | 2.58 |
|  | ≥85 years | 589.3 | 11.2 | 189.4 | 2.82 | 329.3 | 558.9 | 8.33 |

